# Effects of cannabis use on antidepressant treatment response to repetitive transcranial magnetic stimulation and ketamine

**DOI:** 10.1101/2023.06.28.23291446

**Authors:** Mohammad Ali Shenasa, Houtan Totonchi Afshar, Eric A. Miller, Em Ellerman-Tayag, Jyoti Mishra, Dhakshin Ramanathan

## Abstract

**Background:** The antidepressant effects of ketamine and repetitive transcranial magnetic stimulation (rTMS) are hypothesized to rely on mechanisms of long-term-potentiation and synaptic plasticity. Cannabis, via activation of CB1 receptors has been shown to impair synaptic plasticity, suggesting that cannabis use might moderate the antidepressant effects of ketamine and/or rTMS.

**Methods:** We performed a retrospective chart review of 222 Veterans, including 58 females, treated for depression with either rTMS or ketamine/esketamine at the VA San Diego Medical Center (VASDMC). We estimated the effects of treatment using changes in the Patient-Health-Questionnaire 9 (PHQ-9) split by cannabis use. Cannabis use was determined using self-report for rTMS (102 total, 23 screening positive for cannabis use) or urinary drug screens for ketamine (120 total, 40 screening positive for cannabis use). Mixed-level repeated measures ANOVA was utilized to determine whether cannabis use affected PHQ-9 scores (group effect) or the change in PHQ-9 over time (group x time interaction).

**Results:** Cannabis use did not affect overall symptom severity (group effect F (1, 100) = 0.58, p = 0.45) for rTMS, group effect (F (1, 118) = 0.58, p = 0.45) for ketamine, nor did it impact changes in symptoms for either treatment (group x time effect for ketamine: (F (7, 759) = 0.36, p = 0.93); group x time effect for rTMS (F (5, 412) = 0.4160, p = 0.83).

**Conclusions:** Cannabis use was unrelated to antidepressant treatment outcomes for either rTMS or ketamine, suggesting that cannabis use should not be a contraindication for these treatments.

## Introduction

Major depressive disorder (MDD) is a disabling condition with high worldwide prevalence (Whiteford et al., 2013). A sizable proportion of patients with MDD do not respond to traditional first line pharmacotherapy, with a 12-month prevalence of treatment resistant depression (TRD) among patients treated with medication estimated to be 30% (Zhdanava et al., 2021). Several new treatments have been developed for treatment resistant depression in recent years, including repetitive transcranial magnetic stimulation (rTMS) and ketamine. Meta-analyses have found that both rTMS (Brunoni et al., 2017) and ketamine/esketamine (Bahji et al., 2022) are effective for TRD.

The antidepressant effects of both rTMS and ketamine are thought to rely on aspects of synaptic plasticity invoking long-term potentiation (LTP) (Huang et al., 2005; Kim et al., 2023; Wrightson et al., 2023). Thus, agents that affect or diminish synaptic plasticity may likewise reduce the antidepressant effects of ketamine and rTMS. One commonly used agent that impairs synaptic plasticity is cannabis (Hoffman et al., 2021). Cannabis is a widely consumed substance world-wide, with nearly 192 million users in 2018 and an estimated 3.9% of users among the global population aged 15-64 years old (Ransing et al., 2022). Given this prevalence and the potential impacts of cannabis on neural plasticity (Hoffman et al., 2021) and mental health (Moore et al., 2007), we were interested in examining the effects of cannabis use on the antidepressant effects of rTMS and ketamine. To date, there is limited data on the effects of cannabis use on response to either rTMS or ketamine. We hypothesized that cannabis use would impair the antidepressant effects of these treatments. To answer this question, we performed a retrospective analysis of Veterans treated with these modalities at the VA San Diego Medical Center between 2017 – 2023 and analyzed whether cannabis moderated the antidepressant effects.

## Methods

In accordance with the EQUATOR Network, we report our data following STROBE guidelines.

This study was approved as an institutional review board (IRB) exemption by the local VA institutional review board (IRB 1223219). We performed a chart review of patients referred to the neuromodulation clinic at the VA San Diego Medical Center for rTMS or ketamine between January 2017 to March 2023. Veterans were included if they received at least 2 weeks of rTMS and had at least one Patient Health Questionnaire-9 (PHQ-9) post treatment (n = 105), or if they received at least 2 ketamine treatments with at least one PHQ-9 score post-treatment initiation (n= 124). Treatments followed standard clinical protocols. For rTMS treatments, most Veterans received either of the FDA-cleared protocols, 10Hz or intermittent theta-burst stimulation (iTBS), targeted to the left dorsolateral prefrontal cortex (DLPFC). Ketamine/esketamine treatments were offered twice/week for 8 treatments for the induction period. Esketamine dosing typically started at 54mg, and was then flexibly dosed based on clinical response. Ketamine dosing started at 0.5mg/kg and was flexible dosed (up to 1mg/kg) based on clinical response.

Data on cannabis use was gathered during the clinical intake by self-report (for rTMS) or via positive tests on a urinary drug screen (UDS) administered prior to every ketamine treatment. A positive UDS at any point during the 8 ketamine induction treatments was a marker for cannabis use.

### Outcomes

Clinical data used in this study were gathered as part of standard measurement-based care. Our main outcome of interest was change on the PHQ-9, an established self-report questionnaire of the severity of depression symptoms. For rTMS treatments, assessments were given prior to the first treatment and then weekly. For ketamine/esketamine treatments, assessments were given prior to each treatment.

### Statistical analysis

Multilevel mixed-effects model was implemented within Prism to model repeated-measures effects of treatment, a between group factor of cannabis use, and a group x treatment interaction for PHQ-9 symptoms. Statistical analyses were interpreted using an alpha of 0.05, and reported two-sided values. We reported the F statistic (F) for ANOVA analyses and used the Geisser-Greenhouse correction for repeated-measures. Post-hoc power analysis (conducted in G*Power 4) showed that we had 80% power to detect a small difference between groups (f = 0.11).

## Results

Data on age, gender, baseline PHQ-9 data, ketamine modality, and rTMS protocol are included in **Table 1**. To examine group differences by cannabis use, we performed a t-test for normally distributed data and a chi-squared test for dichotomized data. We found a group difference in age for both the rTMS and ketamine groups (cannabis users were on average younger than non-cannabis users (p<0.05). No other group differences were noted.

**Table 1:**
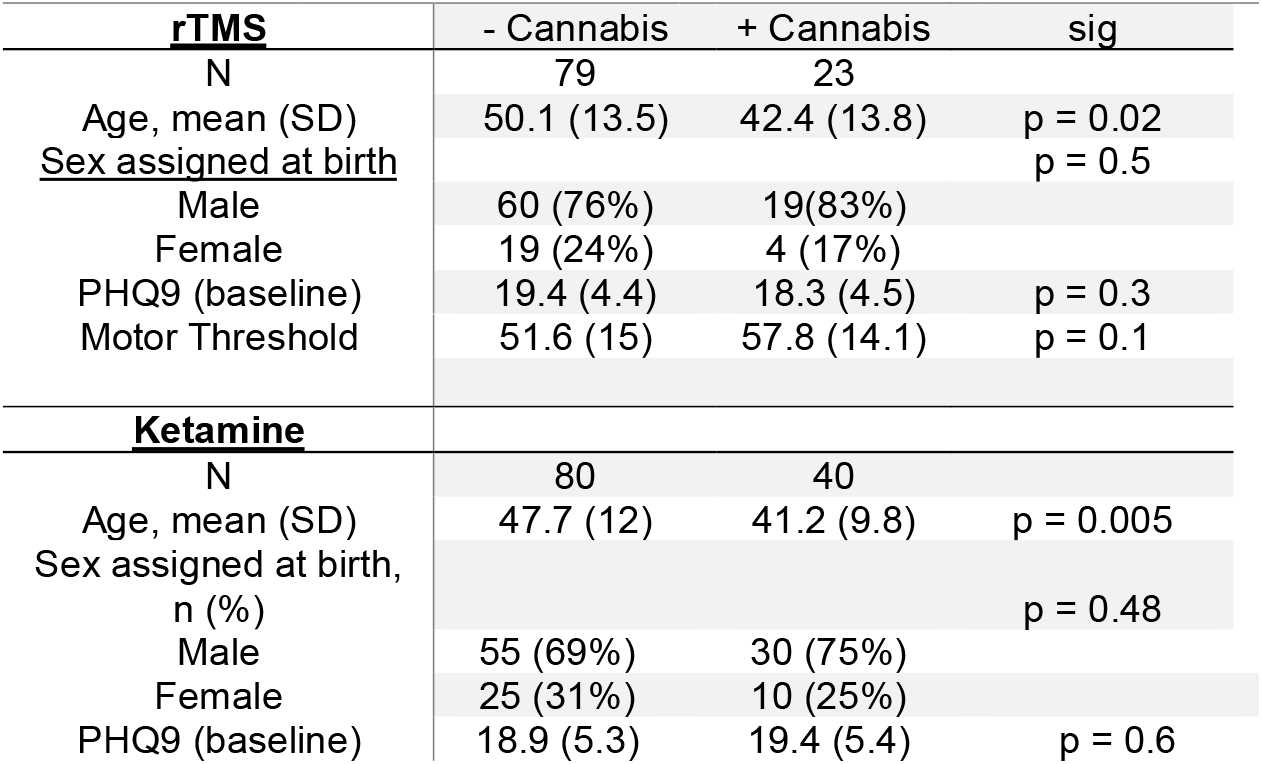
Demographics of subjects split by group. Significant differences by cannabis use were estimated using a one-sample two-sided t-test (age, PHQ-9 scores, MT scores) or a Chi-squared test (sex). Age was significantly different by cannabis use for both the ketamine and rTMS groups.

We next examined whether cannabis use affected changes in PHQ-9 scores during ketamine treatments (**Figure 1**). We found a main effect of treatment (F (4, 510) = 21.6, p<0.0001), with a mean reduction of 4.4 points on the PHQ-9 (CI 2.7-6.1). There was no main effect of cannabis use (F (1, 118) = 0.58, p = 0.45) nor a significant group x time interaction (F (7, 759) = 0.36, p = 0.93). Non-cannabis users showed a 4.5 point reduction (CI 2.539-6.407) while cannabis users showed a 4.3 point reduction (CI 0.7802-7.917). At no time point was there a significant between group difference in PHQ-9 scores (all corrected p values > 0.7).

**Figure 1:**
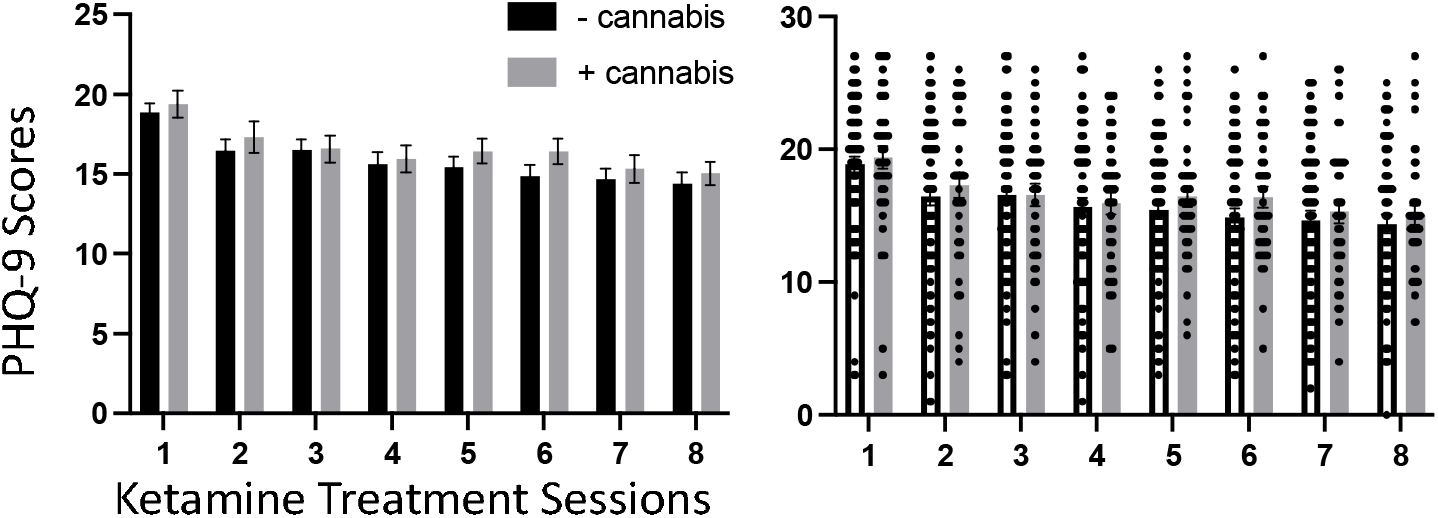
PHQ-9 scores (left panel individual, right panel mean/SEM) separated by cannabis use (UDS). Mean effect across treatments (p<0.0001), but no overall group effect (p = 0.45) or treatment x group effect (p = 0.93), indicating cannabis use does not diminish the antidepressant effects of ketamine.

We next examined whether cannabis use affected treatment outcomes during rTMS treatments (**Figure 2)**. We found a statistically significant effect of treatment on PHQ-9 for rTMS (F (5, 412) = 42.55, p < 0.0001). Across groups there was a 6.5 point reduction in the PHQ-9 (CI 4.9-8.2). There was no main effect of cannabis use on overall PHQ-9 scores (F (1, 100) = 0.58, p = 0.45), nor a group x time interaction (F (5, 412) = 0.4160, p = 0.83). Non-cannabis users showed a 6.9 point reduction (CI 5.40-8.5) while cannabis users showed a 6.1 point reduction (CI 3.2-9), and at no time point was there a significant between group difference in PHQ-9 scores (all corrected p values > 0.7).

**Figure 2:**
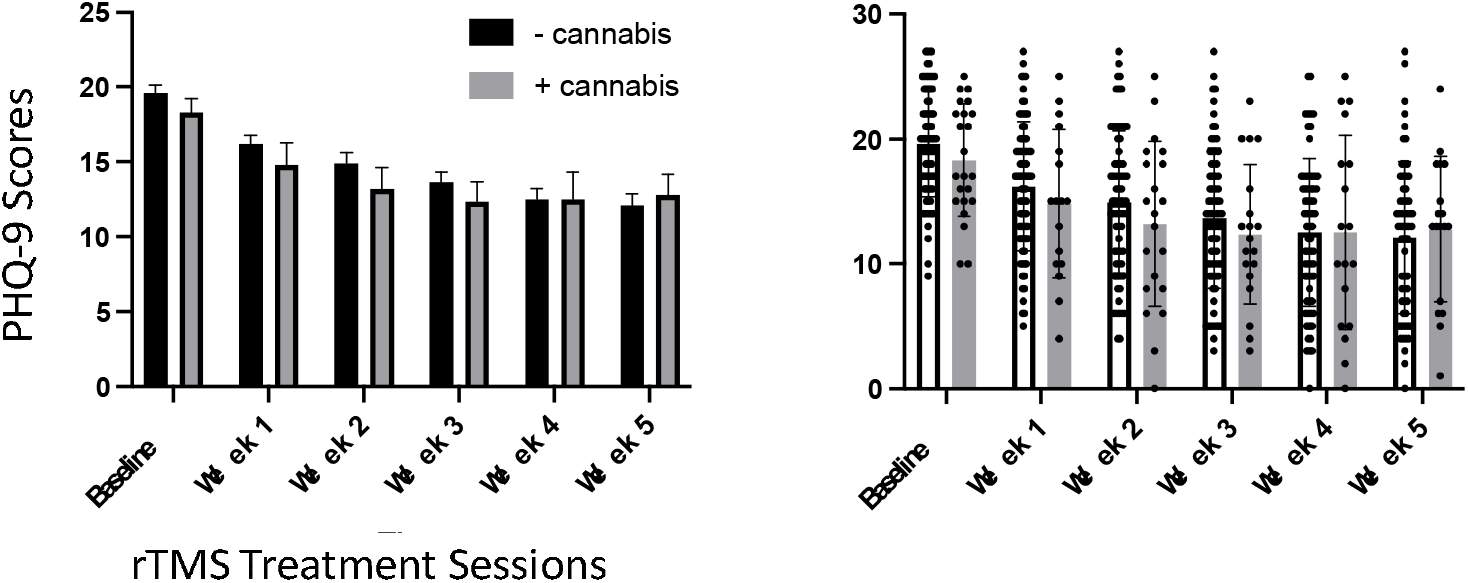
PHQ-9 scores (left panel mean/SEM, right panel individual subject data) separated by cannabis use (self-reported). Main effect of treatment across weeks (p<0.0001), but no overall group effect (p = 0.45) or treatment x group effect (p = 0.84), indicating cannabis use does not diminish the antidepressant effects of rTMS treatments.

Thus, across individuals there was a striking similarity in the lack of effect cannabis use had on the antidepressant efficacy of either ketamine or rTMS therapy.

## Discussion

There is evidence that both ketamine and rTMS act through synaptic plasticity (Huang et al., 2005; Kim et al., 2023) as well as modulation of GABA processes (Lenz & Vlachos, 2016; Voineskos et al., 2021; Zanos & Gould, 2018). Exogenous cannabis use, via action on cannabinoid receptors (particularly the CB1R) (Kendall & Yudowski, 2017) has been shown to both modulate plasticity (Hoffman et al., 2021) and affect cortical inhibition (Fitzgerald et al., 2009), both targets of ketamine/rTMS therapy processes (Huang et al., 2005; Kim et al., 2023; Lenz & Vlachos, 2016; Voineskos et al., 2021; Zanos & Gould, 2018). It was therefore surprising but encouraging that we found no difference in antidepressant response to either ketamine or rTMS based on cannabis use. This suggests that until more data is produced, it is not necessary to require complete abstinence to maximize the effects of these treatments.

There are several limitations to our data. First, this is a modest sample but powered (at 80%) to detect at least a small effect size. Our findings within a unique patient population of Veterans may not generalize to non-Veterans. This analysis was neither randomized nor controlled. Lastly, we did not fully assess the frequency/duration/chronicity of cannabis use, which may be an important determinant. There are however several strengths to this retrospective analysis that render this data informative. First, results generalized across two different treatment modalities (ketamine and rTMS). This study was a naturalistic, real-world study without any obvious bias in terms of recruitment or inclusion/exclusion of cannabis users that might occur in randomized controlled trials. Finally, this is the largest data to date of the effects of cannabis on treatment outcomes of ketamine and rTMS.

## Data Availability

All data produced in the present work are contained in the manuscript

## Role of Funding Source

Funding was received via ORD funding for the Center of Excellence for Stress and Mental Health, and Burroughs-Wellcome Fund Career Award for Medical Scientists on behalf of DR.

## Contributors

MAS and HTA are co-first authors and contributed equally to this work. All authors substantially contributed to this study, manuscript preparation, and approved the final manuscript as submitted.

## Conflict of interest

No authors have disclosures to report.

## Acknowledgements

None.

## Notes

### Competing Interest Statement

The authors have declared no competing interest.

### Author Declarations

This study was approved as an institutional review board (IRB) exemption by the VA San Diego Healthcare system institutional review board (IRB 1223219). Ethical approval was waived for this study.

